# Exploring the impact of shielding advice on the health and wellbeing of individuals identified as extremely vulnerable and advised to shield in Southwest England amid the COVID-19 pandemic: A mixed-methods evaluation

**DOI:** 10.1101/2022.01.05.21268251

**Authors:** Gemma Lasseter, Polly Compston, Charlotte Robin, Helen Lambert, Matthew Hickman, Sarah Denford, Rosy Reynolds, Juan Zhang, Shenghan Cai, Tingting Zhang, Louise E Smith, G James Rubin, Lucy Yardley, Richard Amlôt, Isabel Oliver

## Abstract

**Objective:** Explore the impact and responses to public health advice on the health and wellbeing of individuals identified as clinically extremely vulnerable (CEV) and advised to shield (not leave home for 12 weeks at start of the pandemic) in Southwest England during the first COVID-19 lockdown.

**Design:** Mixed-methods study; structured survey and follow-up semi-structured interviews.

**Setting:** Communities served by Bristol, North Somerset & South Gloucestershire Clinical Commissioning Group.

**Participants:** 204 people (57% female, 54% >69 years, 94% White British, 64% retired) in Southwest England identified as CEV and were advised to shield completed the survey. Thirteen survey respondents participated in follow-up interviews (53% female, 40% >69years, 100% White British, 61% retired).

**Results:** Receipt of ‘official’ communication from NHS England or General Practitioner (GP) was considered by participants as the legitimate start of shielding. 80% of survey responders felt they received all relevant advice needed to shield, yet interviewees criticised the timing of advice and often sought supplementary information. Shielding behaviours were nuanced, adapted to suit personal circumstances, and waned over time. Few interviewees received community support, although food boxes and informal social support were obtained by some. Worrying about COVID-19 was common for survey responders (90%). Since shielding had begun, physical and mental health reportedly worsened for 35% and 42% of survey responders respectively. 21% of survey responders scored ≥10 on the PHQ-9 questionnaire indicating possible depression and 15% scored ≥10 on the GAD-7 questionnaire indicating possible anxiety.

**Conclusions:** This research highlights the difficulties in providing generic messaging that is applicable and appropriate given the diversity of individuals identified as CEV and the importance of sharing tailored and timely advice to inform shielding decisions. Providing messages that reinforce self-determined action and assistance from support services could reduce the negative impact of shielding on mental health and feelings of social isolation.

**Strengths and limitations of this study**

- The mixed-methods study examines the experiences of clinically extremely vulnerable (CEV) people at the height of the COVID-19 crisis, immediately after the first lockdown in England.
- The use of an existing list of individuals identified as needing to “shield” from Bristol, North Somerset & South Gloucestershire (BNSSG) Clinical Commissioning Group (CCG) allowed for access to key patient groups at the height of the crisis.
- Findings may not be applicable to wider CEV populations due to demographic bias.

## Introduction

On 22 March 2020, the Secretary of State for the UK Government announced that individuals in England who, based on understanding at the time faced the highest risk of being hospitalised by COVID-19, should “shield” themselves.^1^ Members of this group were initially advised to not leave their homes for 12 weeks and not go out for shopping, travel, or leisure. This marked the start of what came to be known as ‘shielding’ in England, which was later paused on 1 August 2020.

At the start of the pandemic, the UK Government identified the need to develop a patient list of clinical extremely vulnerable (CEV) people so that they could be sent public health advice and offered support to stay at home and avoid all non-essential contact.^1^ As there was no single mechanism available to support this identification process, a challenging and complex clinical data search was conducted across primary and secondary care settings in England. In total 2.2 million individuals were formally identified as CEV, but various delays were reported in identifying, communicating, and supporting CEV people during this initial period.^1^ For those who were able to officially register with the UK Cabinet Office as CEV,^2^ support with food, medicine, and basic care was offered by central government, local authorities, service providers, charities, rapidly-formed local support groups, neighbours, and relatives.

During the initial 12-week period of shielding over 500,000 people were provided with government funded food parcels.^1^ The advice provided in England suggested that individuals who were shielding should strictly avoid contact with anyone displaying coronavirus symptoms, stay at home, not attend any gatherings, not go out for shopping, leisure or travel, and arrange for food and medication deliveries to be left without social contact. Inside the home, people were advised to minimise time spent with others in shared spaces, keep two metres away from others and sleep in a different bed, use a separate bathroom if possible, and avoid using the kitchen when others were present, eating in a separate room. These are challenging recommendations, and the extent to which they protect people depends largely on the willingness and ability of the population to adhere to them.^3^ Yet it remains unclear whether CEV people fully understood these recommendations, whether they were able to adhere to them, or what impact attempting to adhere to this advice had on their wellbeing. This information would allow us to develop better advice, should such an intervention ever be required again in the future.

The aim of this study was to gain a better understanding from individuals identified as CEV about the effectiveness and acceptability of advice that they received to “shield” during the COVID-19 pandemic, and to explore the reported impact of shielding on their health and wellbeing.

## Methods

A two-stage mixed methods study, including a structured survey and semi-structured telephone interviews with a sample of individuals who had been identified as needing to “shield” by Bristol, North Somerset & South Gloucestershire (BNSSG) Clinical Commissioning Group (CCG).

### Patient and public involvement

Given the extremely rapid and responsive nature of this research, it was not possible to involve patients or the public in the development of the study and associated materials. However, staff at BNSSG CCG were involved in planning the study and facilitating participant recruitment. Additionally, findings from this study will be shared with participants on publication.

### Structured survey

A random sample of 840 people were contacted by post and invited to take part in the structured survey, stratified by index of multiple deprivation (IMD^4^; 240 in the lowest quintile and 150 in each of the remaining quintiles (600 in total)). Potential survey participants were given the option to respond via post or online.

All surveys were completed between 7^th^ August 2020 and 23^rd^ October 2020. The survey consisted of a 54-item questionnaire, including sections on sociodemographic and household characteristics, knowledge of coronavirus symptoms and public health advice, self-reported barriers and facilitators to advice and a self-assessment of mental health and wellbeing. The Patient Health Questionnaire (PHQ-9)^5^ was used to screen for probable depression and the Generalised Anxiety Disorder Scale (GAD-7)^6^ was used to screen for probable anxiety, both using a cut-off point of 10 to indicate the possibility of clinical presentation. The PTSD checklist (PCL-5)^7^ was used to screen for possible post-traumatic stress disorder (PTSD), using a cut-off point of 4 to suggest a potential clinical presentation (full survey available in Supplement 1).

Data from each survey were initially analysed using summary statistics. Not all respondents answered all questions, therefore percentages given below use the number of respondents to each question as a denominator. Where relevant, variables were consolidated into binary indices and compared using a chi-squared test of independence. Statistical analyses were performed in R and RStudio (V1.1.463).^8^

### Qualitative interviews

Interviews were conducted between 26^th^ August and 30^th^ September 2020. At the time of interviews, all four UK nations had relaxed their lockdown measures. Non-CEV member of the general public living in England could return to work if their workplace was considered COVID safe. Non-essential shops and places of worship reopened, but strict social distancing was encouraged (i.e., staying two metres apart). Groups of six individuals from different households were allowed to meet outside. Anyone with COVID-19 symptoms, and their household contacts, were expected to isolate. Most notably, for the purposes of this study, individuals identified as CEV were advised to remain cautious and to stay at home where possible and, if they did go out, to follow strict social distancing.

All responders to the survey were eligible to participate in a qualitative interview. Forty-five respondents consented to take part in a follow-up interview. A convenience sample consisting of those that completed the survey first, consented to participate in the interview, and shared valid contact information was used to identify interview participants. These potential interviewees were initially approached via email and subsequently individuals were emailed an information sheet about the study. All interviews were conducted via telephone by the first author (GL). Participants were offered a £20 shopping voucher as reimbursement for their time.

Interviews lasted between 42 to 69 minutes (median 54). Verbal consent from participants was recorded and a flexible topic guide was used to aid questioning, allowing participants to discuss emerging ideas. Participants were asked about the shielding advice they had received during the first UK lockdown (23^rd^ March 2020 to 1^st^ August 2020), the acceptability of this advice and their resulting behaviours (Supplement 2).

Interviews were transcribed, anonymised, and thematically analysed using NVivo 12 (QSR International).^9^ A subset of transcripts were coded inductively to establish an initial analysis framework (CR). This framework was then applied (GL) while reading each transcript and listening to the interview audio files to help capture verbal emphases. Following initial thematic analysis, two researchers (GL and PC) independently coded a selection of transcripts. Themes relating to participants’ understanding and adherence to the UK Government’s shielding advice were discussed, plus reported experiences and behaviours during the 12-week lockdown. Data were compared to the initial coding framework, with adaptations discussed, agreed, and made as required. All transcripts were subsequently reread and compared numerous times until no new codes were found.^10^ The final coding framework was used to identify themes and descriptive accounts emerging from the data; this approach was used to explore participants’ opinions.^11^

Ethical approval for this study was obtained on 27^th^ May 2020 from the Heath Research Authority and Health and Care Research Wales (Project ID 284629, REC ref 20/HRA/2549).

## Results

### Survey and interview participation

Two hundred and four respondents (of which 117, 58%, female) completed the survey. Most (110, 54%) respondents were over 69 years old, 75 (37%) were between 45 and 69 years old and 18 (9%) were between 25 and 44 years old. One hundred and ninety-one (94%) respondents identified as White British. One hundred and eighty-five respondents gave their occupation: one hundred and twenty-eight (64%) were retired, 22 (11%) working full time, 16 (8%) working part-time, seven (4%) were currently on leave or furloughed, seven (3%) were unemployed and four (2%) were stay at home parents / housemakers.

70% of survey responders lived with their family, 1% shared their property with non-family members and 29% lived alone. Most households contained one (30%) or two (52%) people; 18% lived in a household with more than two people and 14% households contained at least one child under 18. Nearly all survey responders (97%) had access to outside space at home, such as a garden, yard, balcony or terrace.

A total of 13 survey participants (7 female) took part in the interviews. All interviewees were White British, with ages ranging from 25 to over 69 years. Eight interview participants were retired, four were in either full or part-time work and one was a fulltime parent. One interviewee reported having no access to outside space at their home and five participants lived alone.

### Acceptability of official advice to “shield”

Most respondents to the survey (80%) agreed or tended to agree that they received all information required, and most (76%) thought they tried to initially follow all advice related to shielding. People who thought they would try to follow all advice related to shielding initially were more likely to feel that they received all the information they needed (⍰^2^= 7.396, df = 1, *P* = 0.007).

When explored during interview the initial timing of shielding advice was questioned by some participants, especially those that had already begun shielding before they received any official advice (Table 1, Quote 1). Interviewees reported receiving official shielding advice at various times between March and June 2020. Two participants reported never receiving an ‘official letter’ from the government advising them to shield, rather they received confirmation of their CEV status from a variety of sources their healthcare provider or local council. These inconsistencies were problematic for some CEV interview participants, as it meant that they did not receive any formal advice until midway through their shielding period, which ran from 23^rd^ March – 1^st^ August 2020 (Table 1, Quote 2).

**Table 1:** Acceptability of official advice to “shield” to interview participants.

|  |  |
| --- | --- |
| Quote 1 | “I heard on the news in February that this was around, I heard it in January actually, I locked myself in because I knew what was going to happen. So, the actual notification from the NHS was the 23<sup>rd</sup> March at which time I’d already locked myself in for about four weeks at that time” (Shield 5). |
| Quote 2 | “This was the first official notification we had from anybody about [shielding], and it’s dated 22 June. It’s from [local authority organisation] saying that, ‘You were sent a letter from the government as you have been identified as being extremely vulnerable’. Well, the answer to that was, well, no, we didn’t” (Shield 10). |
| Quote 3 | “We did watch the broadcast every day on the television from the government because my husband found it fascinating. I got a bit lost because it’s so complicated...” (Shield 9). |
| Quote 4 | “I started to watch some of the daily briefings from the government and then stopped because I |
|  | was becoming a bit... I thought, oh God, I'll never leave my house ever if I keep watching these! [laughs] So, you know, it became a bit depressing" (Shield 7). |
| Quote 5 | "Watching it on the television sometimes with the announcer like Boris, I'd get confused because they'd say something and then they'd change it" (Shield 9). |
| Quote 6 | "Daily briefings from the government, it almost felt a bit flippant that people like me were told to shield.... I felt a bit as if we were pushed to one side and it's quite fine that your life is put on hold and you have to stay in your house... let's get everything else back to normal but forget about the shielders, they can just stay at home out of the way" (Shield 12). |

After the initial notification to shield, most interview participants noted that they frequently sought additional information to supplement the shielding advice. The daily Government televised briefings were identified as a key information source for all interview participants at the start of the pandemic, although these updates were increasingly supplemented with information from other sources (e.g., radio, newspapers, internet searches, social media, family members, and friends) due to concerns about the comprehensibility, relevance, and consistency of these daily briefings (Quote 3 to 5). When reflecting on the televised briefings a few interview participants talked about the updates and advice not being relevant to shielding CEV individuals and that this led to feelings of being forgotten or disregarded (Quote 6).

### Attitudes and behaviours in response to shielding advice

Two thirds (66%) of the survey responders reported that all people in their household shielded with them by staying at home to avoid contact with others, and further 21% said that other household members tried to shield with them but were not able to. Members of the household were more likely to decide to shield together if all of them were over 70 or CEV (88% compared with 54% if least one person in the household was under 70 and / or not CEV) (⍰^2^= 17.16, df = 1, P < 0.001). Interview participants reported that the shielding advice and perceived risks of COVID-19 infection were considered when deciding on their shielding behaviours, and for some shielding as a household was felt to be the only realistic approach, otherwise they would have been unable to shield in accordance with the government’s advice due to restricted living space or caring responsibilities (Table 2 – Quote 1).

**Table 2:** Interview participants’ attitudes and behaviours in response to the shielding advice

|  |  |
| --- | --- |
| Quote 1 | "The governments advice, to be honest that whilst it was comprehensible, it wasn't necessarily easy to follow. Which is basically, if you were shielding and you're living with somebody else who has not been asked to shield, the advice then went on to say you need to keep two metres distance from them at all times. Ideally sleep in different bedrooms, make different arrangements for your meals, use different toilet facilities etc. etc. And we read this through, and we said well we couldn't actually do that over an extended period of time. It would be absolutely awful you know we're a married couple. We have a shared life; we couldn't be basically isolating from each other within the house for weeks on end... and I said to ((spouse)) just think about it if we're both shielding together there is no need to do that if we are both taking exactly the same precautions, we're not introducing risk into the household... so that was the basis on which we decided largely we were going to shield together" (Shield 3). |
| Quote 2 | "I think you kind of had to follow the guidelines really. I just tweaked them in a bit of a way that I guess suited me at the time. I read the bits that I wanted to read, and I believe I kept to the guidelines as sufficiently as I could for me" (Shield 4). |
| Quote 3 | "As lockdown progressed, I found the urge to go out. I mean I'd be ultra-cautious; I wouldn't go out into crowds; I'd go out at odd hours when nobody was around. I'd wear nitrile gloves and face covering but it was something I had a need to do just to get out from the four walls that sort of it became a bit of a prison I suppose. And I exercised that sort of covert, that sort of outside activity, I felt like I was creeping out put it that way and I did that about once maybe twice a week maximum." (Shield 1) |
| Quote 4 | "We welcomed the fact that we were legitimised to shield so it's had a positive effect. And also it sounds a strange thing to say in a national emergency but in a strange kind of way we're actually certainly for the first couple of months quite enjoyed it because not only was the relief okay we can now sort of stay safe but there was also the fact that right now we've got chance to get on top of all sorts of projects and things that we've been meaning to do for a long time but maybe haven't found time" (Shield 3). |
| Quote 5 | "I don't want to shield again, that's one thing I know, I don't want to do it. I will definitely moderate my behaviour for going out, but I don't want to shield. I don't want anybody to say to me again, you are locked in your home for the foreseeable future" (Shield 12). |

None of the interview participants fully adhered to all the shielding recommendations, indeed a spectrum of shielding adherence behaviours were reported with advice being adapted to suit personal circumstances (Quote 2) and found to wane over time (Quote 3). Key external factors found to impact on the shielding behaviours of participants were access to outside space, food, and medication supplies.

Despite some of the difficulties when following the shielding advice, 71% of survey responders felt that they were likely or highly likely to follow similar shielding advice for another three months if needed. A few interview participants even described the label of “shielding” as socially advantageous, as it legitimised their decision to self-isolate (Quote 4). However, over half of survey responders (56%) thought that it would be hard or very hard to follow such additional restrictions, indeed some interview participants had strong reservations about needing to shield again in the future (Quote 5).

### Impact of “shielding” on health and wellbeing

#### Accessing healthcare

Half of survey responders (51%) had successfully accessed healthcare either virtually or in person since being advised to shield, however an additional 13% experienced some problems. The remaining 36% had not tried to access healthcare. Interviewees also recounted mixed experiences, with some reporting smooth interactions with their healthcare providers, others experiencing initial breakdowns in communication with their GP practice or secondary care specialists that were quickly resolved and a minority having no healthcare interactions during shielding.

Survey responders were presented with a list of possible symptoms that could be attributed to COVID-19. Approximately half of the survey respondents (47%) reported at least one of these symptoms since being advised to shield. The most common signs reported were non-specific: “feeling tired or having low energy” (38%) and “aches and pains” (19%). Approximately a quarter (27%) of people with symptoms sought professional help either on the phone (24%) and /or in person (11%). Assuming that phone calls preceded a visit to a healthcare facility, only two people did not call a healthcare provider prior to visiting a healthcare facility. 42% of survey respondents that reported symptoms modified their behaviour to decrease contact with other people inside and / or outside their household.

#### Impact on physical health

Most survey respondents (81%) did not think, or were sure, they had not had coronavirus, 12% were unsure if they had had it, and 7% thought they had probably or definitely had coronavirus.

Most survey respondents answered that they had a health problem that limited their activities prior to being asked to shield (71%). Despite this, from all survey respondents 67% did not need regular help, 68% did not have health problems that required them to stay at home and 72% did not need to use a stick, walker, or wheelchair to move about (72%). Respondents aged over 69 were more likely to answer yes to at least one of these questions than younger respondents (⍰^2^ = 4.607, df = 1, P = 0.032). More surveyed people thought that shielding was making their physical health worse (10% strongly agreed and 25% tended to agree) than thought shielding was making their physical health better (3% strongly agreed and 5% tended to agree). Furthermore, some interviewees felt that shielding had negatively impacted on their levels of daily exercise (Table 3 - Quote 1).

**Table 3:** Impact of “shielding” on health and wellbeing of interview participants

|  |  |
| --- | --- |
| Quote 1 | “It hasn’t done much for my physical health... and I suppose towards the middle I was, sort of |
|  | losing a bit of motivation... I don't think I was getting the same kind of exercise I was getting before, so that's not great" (Shield 7). |
| Quote 2 | "I think when you're stuck in the house and someone has told you that you cannot leave your home, you feel a bit trapped. I am really lucky, I have quite a big house, I've got a lovely garden and we had nice weather so I could go outside but it's just not the same as going out and interacting with other people and just getting a bit of fresh air... When you are told for a really prolonged period that you've got to stay at home, it does make you feel like a caged animal because you're trapped in the house. You can't speak to anybody, you can't go out or do anything." (Shield 13) |
| Quote 3 | "I couldn't practically shield from [our children] but they still needed to have regular exercise and be outside, so that caused some anxiety." (Shield 13) |
| Quote 4 | "I'd probably have another course of medication to treat the mental illness I could have suffered and I'm not exaggerating that one I don't think. No, I would have gone you know it was bordering on the crazy I think just being locked up for three months in isolation it wasn't going to be for me." (Shield 1) |
| Quote 5 | "I seemed to get more emotional. I tend to worry a lot and then when you watch the news and you see people in – and I think, why am I worrying, because they've got a far worse deal than me? So, I've got to sort my head out." (Shield 9) |
| Quote 6 | ...as someone shielding, some of the advice – especially as things started to relax a little for the general population – it became a bit depressing actually." (Shield 8) |
| Quote 7 | "The government's bumbled along and hasn't really got a grip on it and we've got a load of the public now thinking they can make it up as they go along as well and that is anxiety provoking for people in the shielding group." (Shield 3) |

33% survey respondents strongly agreed (14%) or tended to agree (19%) that they had enjoyed spending more time at home while shielding. The availability of private gardens and outside spaces was mentioned by interviewees as a key resource for exercise and leisure throughout the shielding period, nevertheless some individuals still reported feeling trapped due to impact of shielding on their sense of freedom (Quote 2). An added complication for some interviewed parents was ensuring their children received sufficient exercise, while also personally maintaining a shielding status (Quote 3).

#### Impact on mental health

180 (90%) respondents to the survey described themselves as “somewhat”, “very” or “extremely” worried about coronavirus. Older respondents tended to be less worried about coronavirus compared with younger respondents (86% of respondents over 69 years compared with 96% of those 69 years or under were at least somewhat worried about coronavirus; ⍰^2^= 4.68, df = 1, P = 0.030).

More survey responders thought that being asked to shield had made their mental health worse (14% strongly agreed and 28% tended to agree) than thought that shielding was making their mental health better (2% strongly agreed and 4% tended to agree). When explored during interviews, various participants reported heightened emotions, worry and depression, and linked these emotions to feelings of being “locked up” for a prolonged periods of time and anxiety about the future as CEV individuals (Quotes 4 to 7).

#### Depression and anxiety

21% of survey respondents had a score of 10 or above on the PHQ-9 questionnaire, indicating possible depression where treatment may be recommended (**Error! Reference source not found**.). 15% respondents scored 10 or above on the GAD-7 questionnaire, indicating a level of anxiety where treatment may be recommended (**Error! Reference source not found**.). Respondents with a score of 10 or over were more likely to report that their mental health had declined since shielding had begun (76%;). For both groups respondents were more likely to report that mental health had declined since shielding had begun; 76% of responders with a PHQ-7 score of 10 or above (⍰^2^= 21.314, df = 1, P <0.00001) and 72% of responders with a GAD-7 score of 10 or above(⍰^2^= 9.863, df = 1, P = 0.0017). 4% of survey respondents had a score of four or more on the PTSD-4 scale, suggesting they could have high likelihood of developing PTSD as a result of their experience of shielding. 28% of survey respondents had felt somewhat (19%), moderately (8%) or very (2%) angry about being told to shield, although these feelings were not discussed by interview participants.

## Discussion

Early in the pandemic the importance of providing clear, tailored advice for patients who were required to shield, alongside appropriate support, was identified.^12-14^ Although findings from this study showed that official shielding advice offered to CEV individuals during the first lockdown in England was deemed to be sufficient by 80% of survey participants, interviewees criticised the delayed timing of this advice and frequently sought supplementary information to inform shielding behaviours. The individual focus of shielding advice was considered impractical and restrictive by some participants, with 66% of survey responders considering it necessary to shield with all household contacts. Interview participants described a spectrum of rational adaptations to the advice, with adjustments based on living situation and personal perceptions of risk.^15^ Few people reported receiving community support. This source of assistance could be strengthened in the future, potentially through involving pre-existing community-based organisations,^16^ charities, volunteer organisations, and / or faith-based institutions.^17^ These groups could also provide a mechanism for disseminating updated advice and maintaining contact with people who are shielding. Accessing such support networks has been suggested as a protective measure against isolation and emotional distress by promoting feelings of social connection,^18^ which may help reduce symptoms of anxiety or depression.^19-21^

Being formally identified as a “shielding” individual was considered socially advantageous by some interviewees as it legitimised their socially avoidant behaviours. But for others this approach resulted in feeling “othered” as a CEV individual,^22^ an issue that was further exacerbated by televised government briefings that lacked CEV specific advice, while providing reassurance to the general public that COVID-19 was most severe in those with underlying health conditions.^23 24^ Social isolation caused by the response to the COVID-19 pandemic has been shown to have a negative influence on mental health parameters.^21 25^ Of the surveyed CEV individuals, 90% were worried about COVID, with 35% agreeing that shielding was making their physical health worse and 43% reporting a negative impact on their mental health, which may have compounded these feelings of social isolation. This emphasises the importance of using communication approaches in the future that avoid implied or unintentional stigmatising of any ‘vulnerable’ group; instead providing messages framed for target groups, and that are identity affirming, promote social unity, and delivered by the right people.^26^

This study was able to integrate quantitative and qualitative information to triangulate information from patients identified as CEV. However, our study population does not represent the general population, as 94% identified as White British, 64% were retired and 54% were aged over 69 years. It is therefore possible that our findings may not be applicable to wider CEV populations due to demographic and experience bias. Our study may also have been influenced by response bias, especially for interviewees who may have been more likely to want to discuss their shielding experiences. As discussed by Timmons et al (2021), it is possible that participants were unable to accurately recall or measure their compliance with COVID-19 shielding advice. These limitations should be considered when transferring the results of this study to other populations, or when using these finding to inform future COVID-19 public health policies.

Individuals who are CEV are a clinically heterogenous group. For the first COVID-19 lockdown in England this contributed to the difficulties in disseminating shielding advice as well as differing adaptations to that advice. This has been highlighted by other authors, in particular that some more vulnerable patients did not receive appropriate advice in time.^27^ Future communications should be disseminated promptly, tailored to health conditions, and delivered in a more targeted way with integrated support from healthcare and mental health services. Furthermore, additional work could focus on future adherence to shielding advice and long-term social distancing adaptations, as well as the long-term implications of shielding for mental health and feelings of social identity. Additionally, creating a mechanism for clear communication, integrated with clinical needs, would support patients who are CEV who may need to shield in future pandemics.

## Data Availability

All data produced in the present study are available upon reasonable request to the authors

## Original protocol

Document attached

## Author’s contributions

All authors except P.C. conceptualised and designed the study. All authors were involved in interpretation of the study’s results. GL, PC, and CR were involved in data analysis. GL & PC led the drafting of the manuscript. All authors reviewed the manuscript, approved the final content, and met authorship criteria.

## Availability of data and materials

The datasets used and / or analysed during the current study are available from the corresponding author on reasonable request.

## Consent for publication

All participants provided written or oral consent for data to be included in publications

## Acknowledgements

G.L., P.C., H.L., M.H., S.D., I.O., C.R., R.R. and L.Y. are supported by the NIHR Health Protection Research Unit (HPRU) in Behavioural Science and Evaluation at the University of Bristol in partnership with UK Health Security Agency (UK HSA).

L.S. and J.R. are supported by the NIHR HPRU in Emergency Preparedness and Response at King’s College London in partnership with UK HSA.

L.Y. is an NIHR Senior Investigator and her research programme is partly supported by NIHR Applied Research Collaboration (ARC)-West, NIHR Health Protection Research Unit (HPRU) in Behavioural Science and Evaluation, and the NIHR Southampton Biomedical Research Centre (BRC).

C.R. is affiliated to the National Institute for Health Research Health Protection Research Unit (NIHR HPRU) in Emerging and Zoonotic Infections at the University of Liverpool in partnership with UK HSA in collaboration with the Liverpool School of Tropical Medicine and The University of Oxford, the NIHR HPRU in Gastrointestinal Infections at the University of Liverpool in partnership with UK HSA, in collaboration with the University of Warwick and the NIHR HPRU in Behavioural Science and Evaluation at the University of Bristol, in partnership with UK HSA. C.R. is based at UK HSA.

The views expressed are those of the authors and not necessarily those of the NIHR, the Department of Health and Social Care or UK HSA. The funders had no role in the design of the study, collection, analysis and interpretation of the data, or in writing the manuscript.

