## Supplementary material for "Exploring the impact of shielding advice on the health and wellbeing of individuals identified as extremely vulnerable and advised to shield in Southwest England amid the COVID-19 pandemic: A mixed-methods evaluation": Figure 1

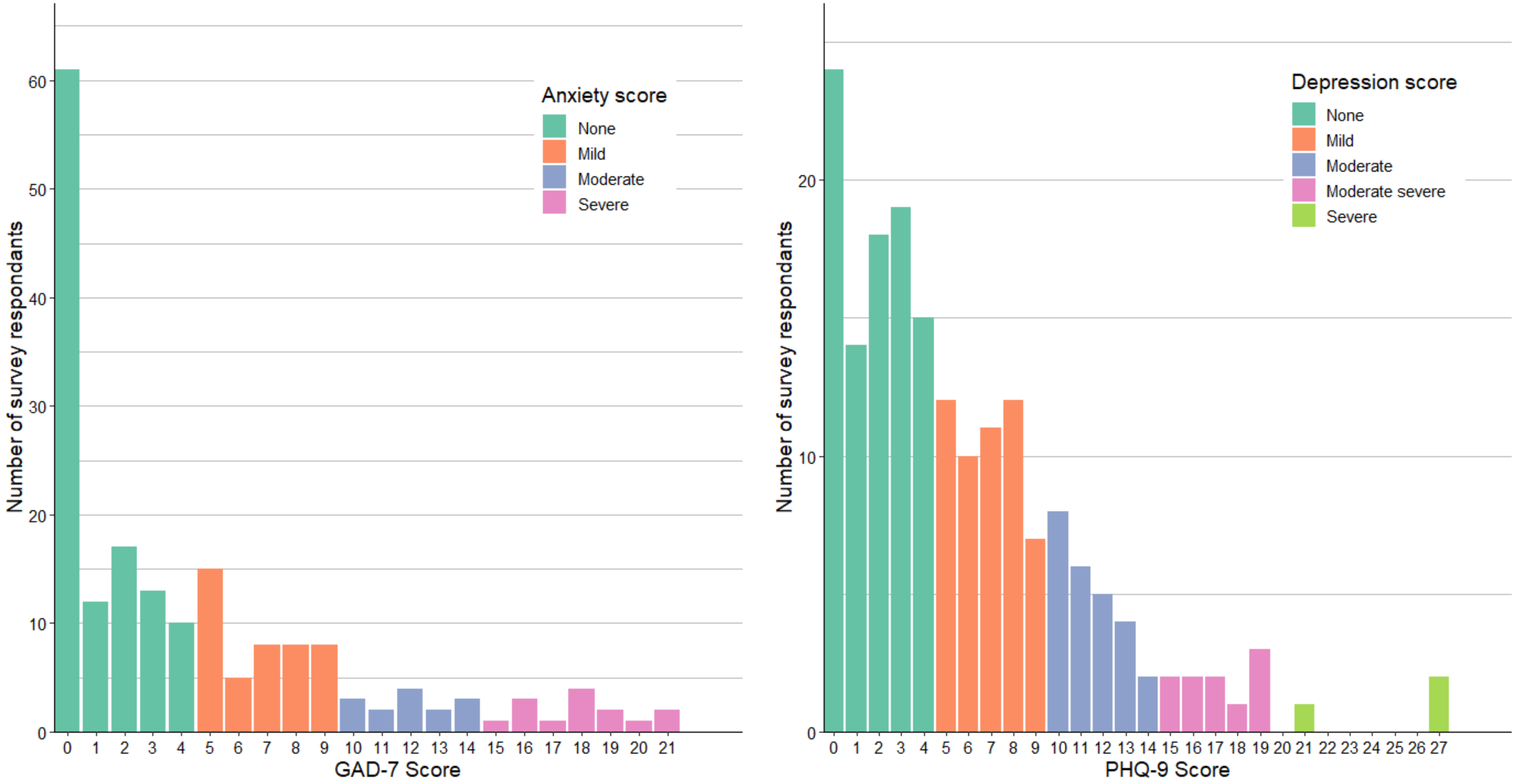


**Figure 1: Survey results for the (A) GAD-7 and (B) PHQ-9 questionnaires** (Categories indicated in the key are as follows. For the GAD-7 cut-off values used: None = 0-4; Mild = 5-9; Moderate = 10-14; Severe = 15-21. For the PHQ-9 cut-off values used were: None = 0-4; Mild = 5-9; Moderate = 10-14; Moderate severe = 15-19; Severe = 15-19. For both questionnaires a cut-off of 10 was used to indicate possible anxiety or depression, respectively.)
