## Supplement 1 for "Exploring the impact of shielding advice on the health and wellbeing of individuals identified as extremely vulnerable and advised to shield in Southwest England amid the COVID-19 pandemic: A mixed-methods evaluation"

### Views on advice to "shield" during the coronavirus (COVID-19) outbreak

Thank you for completing this survey. The information you provide will help us provide support for other people who are "shielding". Please ensure you have read the enclosed information sheet and completed the consent form before completing the survey.

Q1 Overall, how worried are you about coronavirus? (Please select the option that BEST applies to you)

- ☐ Extremely worried
- ☐ Very worried
- ☐ Somewhat worried
- ☐ Not very worried
- ☐ Not at all worried
- ☐ Don't know

Q2 Thinking about before the coronavirus outbreak, for each of the following questions. (Please select one option on each row)

| | Yes | No |
|---|---|---|
| In general, did you have health problems that required you to limit your activities? | ☐ | ☐ |
| Did you need someone to help you on a regular basis? | ☐ | ☐ |
| In general, did you have health problems that required you to stay at home? | ☐ | ☐ |
| If you needed help, could you count on someone close to you? | ☐ | ☐ |
| Did you regularly use a stick, walker or wheelchair to move about? | ☐ | ☐ |

Q3 In general, before the coronavirus outbreak, did you **need support** with getting essential supplies and basic care? (Please tick all that apply)

- ☐ Yes, from my family
- ☐ Yes, from my friends
- ☐ Yes, from my neighbours
- ☐ Yes, from community support (e.g. local volunteers or Facebook group)
- ☐ Yes, from an organisation (e.g. NHS, local council, Government, charity)
- ☐ I did not need any additional support
- ☐ Other

If other, please provide details:

### Advice to "shield"

The following questions refer to when you were contacted by your GP or NHS England because you have a specific medical condition, which increases the risk of severe illness from coronavirus. You were strongly advised to stay at home at all times and minimise fact-to-face contact with other people, this is called “shielding”. Shielding means:

1. Do not leave your home.
2. Do not attend any gathering, including with friends and family in private spaces, for example family homes.
3. Do not have any visitors to your home, except essential carers.
4.Strictly avoid contact with someone who is displaying symptoms of coronavirus.

The Government initially advised people to shield for 12 weeks (until the end of June) and is regularly monitoring this position.

Q4 When I was advised to “shield”, I felt I received all the information I needed. (Please select the option that BEST applies to you)

- ☐ Strongly agree
- ☐ Tend to agree
- ☐ Neither agree or disagree
- ☐ Tend to disagree
- ☐ Strongly disagree

Q5 When I was advised to “shield”, I initially decided I would… (Please select the option that BEST applies to you)

- ☐ Try to follow the advice in full
- ☐ Try to follow the advice, but not completely
- ☐ Not try to follow it at all

Q6 Since you were advised to “shield”, have you **asked for support** with getting essential supplies and basic care? (Please tick all that apply)

- ☐ Yes, from my family
- ☐ Yes, from my friends
- ☐ Yes, from my neighbours
- ☐ Yes, from community support (e.g. local volunteers or Facebook group)
- ☐ Yes, from an organisation (e.g. NHS, local council, Government, charity)
- ☐ I have not asked for any additional support
- ☐ Other

If other, please provide details:

Q7 Since you were advised to “shield”, have you **received support** with getting essential supplies and basic care? (Please tick all that apply)

- [ ] Yes, from my family
- [ ] Yes, from my friends
- [ ] Yes, from my neighbours
- [ ] Yes, from community support (e.g. local volunteers or Facebook group)
- [ ] Yes, from an organisation (e.g. NHS, local council, Government, charity)
- [ ] I have not received any additional support
- [ ] Other (please provide details)

If other, please provide details:

Q8 Since being advised to “shield”, have you experienced any of the following symptoms? (Please tick all that apply)

- [ ] A new continuous cough — If you tick yes to a new continuous cough, please also answer Q9
- [ ] A high temperature/fever
- [ ] Shortness of breath/difficulties breathing
- [ ] Runny or blocked nose
- [ ] Aches and pains
- [ ] Chest pains
- [ ] Chills/shivering
- [ ] Sore throat
- [ ] Diarrhoea
- [ ] Headache
- [ ] Stomach ache
- [ ] Feeling tired or having low engergy
- [ ] Loss of sense of smell or taste
- [ ] None of these — If you have not experienced any of these symptoms, please go to Q11
- [ ] Don't know

Q9 You reported a new continuous cough. Was this a: (Please select one option)

- [ ] Dry cough
- [ ] Wet cough (with mucus or phlegm)
- [ ] Don't know

Q10 When your symptoms developed, what did you do? (Please tick all that apply)

- ☐ Waited a day or two to see if my symptoms got better, before making any changes
- ☐ Checked what to do online
- ☐ Called NHS 111
- ☐ Called 999
- ☐ Went in person to a doctor's surgery
- ☐ Made a phone call to a doctor's surgery
- ☐ Went to a walk-in centre
- ☐ Went to a hospital's Accident and Emergency department
- ☐ Went to a pharmacist/chemist
- ☐ Stayed at home, not leaving home for any reason
- ☐ Reduced the number of times I went out
- ☐ Avoided contact with people inside my household
- ☐ Avoided contact with people outside my household
- ☐ Asked friends or family for advice
- ☐ Did nothing
- ☐ Don't know
- ☐ Something else

If something else, please provide details:

Q11 Do you think you have had coronavirus? (Please select the option that BEST applies to you)

- ☐ I have definitely had it
- ☐ I think I have probably had it
- ☐ I don't know if I have had it or not
- ☐ I don't think I have had it
- ☐ I have definitely not had it

Q12 Have you had a test for coronavirus?

- ☐ Yes, test said I had coronavirus
- ☐ Yes, test said I didn't have coronavirus
- ☐ No, never been tested

### Household characteristics

*The following questions are about where you are living while you are "shielding". If you have moved since being advised to "shield", please respond to these questions for where you will be spending the majority of your time while "shielding".*

Q13 How many people live in your household (including you)? If you were living alone, please write "1".

Q14 How many people in each age group do you live with (not including you)? Please write 0 in any box for which there is no one in your household in that age group.

Children aged 0-4 years ☐

Children aged 5-17 years ☐

Adults aged 18-69 years ☐

Adults aged 70 years and over ☐

Q15 Which of the following best describes your living situation? (Please select one answer)

- ☐ I live by myself — If you live by yourself, go to Q18
- ☐ I live with family (this includes your partner, children or other family by birth or marriage)
- ☐ I live in shared accommodation
- ☐ Other

If other, please provide details:

☐

Q16 Do you share bathroom facilities at home?

- ☐ Yes
- ☐ No

Q17 Do you have a room in your home that you could live and sleep in without coming into contact with other people?

- ☐ Yes
- ☐ No

Q18 Does your home include access to any outside space, such as a garden, yard, balcony or terrace?

- ☐ Yes
- ☐ No

Q19 Do you have any pets that lived in your home?

- ☐ Yes
- ☐ No — If you do not have pets, go to Q21

Q20 Please provide details of pets that lived in your home: (Please select one option on each row)

| | Yes | No |
|---|---|---|
| Dog(s) | ☐ | ☐ |
| Cat(s) | ☐ | ☐ |
| Bird(s) | ☐ | ☐ |
| Reptile(s) | ☐ | ☐ |
| Other | ☐ | ☐ |

If other, please provide details:

#### Experiences of “shielding”

*The following questions are about your experiences since being contacted by NHS England and advised to “shield”.*

*Please remember that your answers are always treated confidentially and are never analysed individually.*

Q21 Since being contacted by NHS England and advised to “shield”, did other people in your household stay at home and avoid contact with other people (i.e. "shield" with you)? (Please select the option that BEST applies to you)

- ☐ Yes
- ☐ No
- ☐ They tried, but were not able to
- ☐ Not sure
- ☐ N/A - I live by myself

Q22 Since being contacted by NHS England and advised to “shield”, how often did you leave your home for each of the following reasons? (Please select one option on each row)

| | Not at all | Occasionally | More than half the days | Nearly every day | Not applicable |
|---|---|---|---|---|---|
| To go to the shops for groceries, toiletries or medicines | ☐ | ☐ | ☐ | ☐ | ☐ |
| To go to the shops for other items | ☐ | ☐ | ☐ | ☐ | ☐ |
| For exercise | ☐ | ☐ | ☐ | ☐ | ☐ |
| For a medical need (e.g. an outpatient appointment) | ☐ | ☐ | ☐ | ☐ | ☐ |
| To go to work | ☐ | ☐ | ☐ | ☐ | ☐ |
| To take my children to or from school/day care | ☐ | ☐ | ☐ | ☐ | ☐ |
| To provide help to someone else | ☐ | ☐ | ☐ | ☐ | ☐ |
| To meet friends or family members who don't live with you | ☐ | ☐ | ☐ | ☐ | ☐ |
| To walk my dog | ☐ | ☐ | ☐ | ☐ | ☐ |
| For another reason | ☐ | ☐ | ☐ | ☐ | ☐ |

If for another reason, please provide details:

Q23 If you left home during your isolation period, how much time did you spend indoors with other people, but keeping 2 metres away from them?

- [ ] No time
- [ ] Less than 15 minutes
- [ ] Less than one hour
- [ ] Several hours
- [ ] One or more days
- [ ] N/A - I did not leave my home

Q24 If you left your home during your isolation period, how much time did you spend indoors with other people and closer than 2 metres from them?

- [ ] No time
- [ ] Less than 15 minutes
- [ ] Less than one hour
- [ ] Several hours
- [ ] One or more days
- [ ] N/A - I did not leave my home

Q25 If you left your home during your isolation period, how often did you have to touch any surfaces other people had touched (for example, to open doors or pay for things)?

- [ ] Never
- [ ] A few times
- [ ] Many times
- [ ] N/A - I did not leave my home

Q26 Since being advised to “shield”, if you have tried to get online groceries delivered, have you been able to? (Please select the option that BEST applies to you)

- [ ] Yes
- [ ] I tried, but was unable to access the website/book a delivery slot
- [ ] I did not try to get groceries delivered

Q27 Since being advised to “shield”, have you had any problems accessing healthcare? (Please select one option)

- [ ] I accessed healthcare but did not experience any problems
- [ ] I accessed healthcare and did experience problems
- [ ] I have not tried to access healthcare

If you have not tried to access healthcare, go to Q29

Q28 Since being advised to “shield”, how often have you had problems accessing healthcare? (Please select one option)

- [ ] Occasionally when trying to access healthcare
- [ ] More than half the times I tried to access healthcare
- [ ] Nearly every time I tried to access healthcare

Q29 Since being advised to “shield”, have you had any visitors in your home? (Please select the option that BEST applies to you)

- ☐ Not at all
- ☐ Occasionally
- ☐ More than half the days
- ☐ Nearly every day

Q30 Since being advised to “shield”, how much of the time have you stayed in your room with the door closed, only coming out when necessary (e.g. to use the bathroom or prepare food)? (Please select the option that BEST applies to you)

- ☐ Nearly every day
- ☐ More than half the days
- ☐ Occasionally
- ☐ Not at all
- ☐ NA - I live by myself

Q31 Since being advised to “shield”, did any of these reasons make it difficult for you to "shield" at home (this means staying on your own in your room with the door closed, only coming out when necessary (e.g. to use the bathroom or prepare food)? (Please select all that apply)

- ☐ There was no room I could use to stay in on my own
- ☐ I had to look after other people (e.g. children, old or sick family members)
- ☐ Other family members wanted or needed to talk to or see me
- ☐ I was very ill and so family members had to come in my room to look after me
- ☐ None of these applied to me
- ☐ Other

If other, please provide details

Q32 Since being advised to “shield”, did you use a separate bathroom or set up a bathroom rota?

- ☐ Yes
- ☐ No
- ☐ N/A - I live by myself

Q33 Since being advised to “shield”, did you share hand towels or kitchen equipment with other people you were living with?

- ☐ Yes
- ☐ No
- ☐ NA - I live by myself

Q34 At the time you were contacted and advised to “shield”, did you do any paid or voluntary work outside your home? (Please select all that apply)

- ☐ I was self-employed, working outside my home
- ☐ I was in full-time paid work outside my home
- ☐ I was in part-time paid work outside my home
- ☐ I was in full-time education and studying outside my home
- ☐ I was in part-time education and studying outside my home
- ☐ I was taking part in voluntary work outside my home
- ☐ I was helping to care for someone outside my home (e.g. a friend or relative)
- ☐ None of these applied to me

If none of these applied to you, go to Q37

Q35 Since being advised to “shield”, did you continue to work outside your home?

- ☐ Yes
- ☐ No

Q36 Why did you continue to work outside your home? (Please select all that apply)

- ☐ I worked as a key/critical worker
- ☐ I could not afford to stop working
- ☐ I was worried about losing my job
- ☐ My employer asked me to go into work
- ☐ I could not do my job from home
- ☐ I had an important task that I needed to do in person (e.g. tend to animals, work in a lab etc.)
- ☐ I wanted to reduce the workload of my colleagues
- ☐ I needed to go into work to sustain my business
- ☐ Other

If other, please provide details:

Q37 Thinking about the advice to "shield", please tell us to what extent you agree or disagree with each of the following statements. (please tick one box for each statement)

| | Strongly agree | Tend to agree | Neither agree or disagree | Tend to disagree | Strongly disagree |
|---|---|---|---|---|---|
| If I had completely followed the advice to "shield", I would have lost touch with my friends and family | ☐ | ☐ | ☐ | ☐ | ☐ |
| My friends or family would have disapproved if I had not completely followed the advice to "shield" | ☐ | ☐ | ☐ | ☐ | ☐ |
| If I didn't completely follow the advice to "shield", I could have been in trouble with the police | ☐ | ☐ | ☐ | ☐ | ☐ |
| If I completely followed the advice to "shield", it would have helped save lives | ☐ | ☐ | ☐ | ☐ | ☐ |
| If I completely followed the advice to "shield", it would have helped protect the NHS | ☐ | ☐ | ☐ | ☐ | ☐ |
| If I had caught coronavirus, I may have become very ill | ☐ | ☐ | ☐ | ☐ | ☐ |
| If I had caught coronavirus, it would have had a severe impact on my family's wellbeing | ☐ | ☐ | ☐ | ☐ | ☐ |
| If I had completely followed the advice to "shield" there would have been more conflict with the people that I was living with | ☐ | ☐ | ☐ | ☐ | ☐ |
| If I had left home and met other people, I could have passed coronavirus to someone | ☐ | ☐ | ☐ | ☐ | ☐ |
| If I leave home and meet other people, I could catch coronavirus | ☐ | ☐ | ☐ | ☐ | ☐ |
| Completely following the advice to "shield" would have had a negative impact on how much money I have | ☐ | ☐ | ☐ | ☐ | ☐ |
| If I completely follow the advice to "shield" I would not be able to carry out important religious activities | ☐ | ☐ | ☐ | ☐ | ☐ |
| While I am "shielding", I am receiving help from someone outside my household | ☐ | ☐ | ☐ | ☐ | ☐ |
| "Shielding" is making my physical health worse | ☐ | ☐ | ☐ | ☐ | ☐ |
| "Shielding" is making my mental health worse | ☐ | ☐ | ☐ | ☐ | ☐ |
| "Shielding" is making my physical health better | ☐ | ☐ | ☐ | ☐ | ☐ |
| "Shielding" is making my mental health better | ☐ | ☐ | ☐ | ☐ | ☐ |
| I enjoy spending more time at home while I "shield" | ☐ | ☐ | ☐ | ☐ | ☐ |

Q38 If you are advised to “shield” for another three months, how likely do you think you would be to follow this advice? (Please select the option that BEST applies to you)

- ☐ Highly unlikely
- ☐ Unlikely
- ☐ Neither unlikely or likely
- ☐ Likely
- ☐ Highly likely

Q39 How hard would it be to follow this advice? (Please select the option that BEST applies to you)

- ☐ Very hard
- ☐ Hard
- ☐ Neither hard or easy
- ☐ Easy
- ☐ Very Easy

Q40 Thinking about people in England who have also been advised to "shield", what percentage do you think are fully following the NHS's recommendations to "shield"? Please enter a number between 0 and 100

☐

Q41 Since being advised to “shield”, how often on average did you… (Please tick one box on each row)

| | Nearly every time | More than half the time | Occasionally | Not at all |
|---|---|---|---|---|
| wash your hands with soap and water, for more than 20 seconds | ☐ | ☐ | ☐ | ☐ |
| clean objects or surfaces that you have touched | ☐ | ☐ | ☐ | ☐ |
| keep 2 metres away from people you live with | ☐ | ☐ | ☐ | ☐ |

Q42 Since being advised to “shield”, have you… (Please select one option on each row)

| | Yes | No | NA |
|---|---|---|---|
| avoid contact with your pet(s) | ☐ | ☐ | ☐ |
| wash hands immediately before and after coming into contact with your pet(s) | ☐ | ☐ | ☐ |
| arrange for someone else to help care for your pet(s) (e.g. dog walking) | ☐ | ☐ | ☐ |

Q43 Thinking about going out in public in the current situation (i.e. not including any healthcare, key work, etc.), to what extent do you agree or disagree with the following statements about wearing masks? (Please tick one box for each statement)

| | Strongly Agree | Tend to agree | Neither agree or disagree | Tend to disagree | Strongly disagree |
|---|---|---|---|---|---|
| Wearing a mask outside of the home can help to protect me from getting coronavirus | ☐ | ☐ | ☐ | ☐ | ☐ |
| Wearing a mask outside of the home can help to prevent me infecting other people with coronavirus if I happen to have it | ☐ | ☐ | ☐ | ☐ | ☐ |
| Only health and other key workers should wear masks, even if there is an unlimited supply | ☐ | ☐ | ☐ | ☐ | ☐ |
| The Government should advise everyone in the UK to wear masks outside of the home | ☐ | ☐ | ☐ | ☐ | ☐ |
| The Government should ban wearing masks unless you are a health or other key worker, even if there is an unlimited supply | ☐ | ☐ | ☐ | ☐ | ☐ |
| I would feel safer going outside if I wear a mask | ☐ | ☐ | ☐ | ☐ | ☐ |
| I would feel more vulnerable going outside if I wear a mask | ☐ | ☐ | ☐ | ☐ | ☐ |
| I dislike people wearing masks because I cannot see their expression | ☐ | ☐ | ☐ | ☐ | ☐ |
| I feel suspicious of people wearing masks because they may have coronavirus | ☐ | ☐ | ☐ | ☐ | ☐ |

Q44 To what extent do you agree or disagree with the following statements? (Please tick one box for each statement)

| | Strongly agree | Tend to agree | Neither agree or disagree | Tend to disagree | Strongly disagree |
|---|---|---|---|---|---|
| If I had coronavirus I would be willing for the data from my mobile phone to be used by public health doctors to identify the places I had visited in the past 7 days | ☐ | ☐ | ☐ | ☐ | ☐ |
| There should be a law that allows public health doctors to access the mobile phone data of people with coronavirus to identify the places they had visited in the past 7 days | ☐ | ☐ | ☐ | ☐ | ☐ |
| During a national emergency we should be less concerned about the privacy of our data | ☐ | ☐ | ☐ | ☐ | ☐ |

### How have events affected you?

The following questions all relate to how recent events might have affected you.

Q45 Thinking over the **last two weeks**, how often have you been bothered by the following problems? (Please tick one box for each row)

| | Not at all | Several days | More than half the days | Nearly every day |
|---|---|---|---|---|
| Feeling down, depressed, or hopeless | ☐ | ☐ | ☐ | ☐ |
| Little interest or pleasure in doing things | ☐ | ☐ | ☐ | ☐ |
| Feeling tired or having no energy | ☐ | ☐ | ☐ | ☐ |
| Trouble falling or staying asleep, or sleeping too much | ☐ | ☐ | ☐ | ☐ |
| Poor appetite or overeating | ☐ | ☐ | ☐ | ☐ |
| Feeling bad about yourself – or that you are a failure or have let yourself or your family down | ☐ | ☐ | ☐ | ☐ |
| Trouble concentrating on things, such as reading the newspaper or watching television | ☐ | ☐ | ☐ | ☐ |
| Moving or speaking so slowly that other people have noticed? Or the opposite, being so fidgety or restless that you have been moving around a lot more than usual? | ☐ | ☐ | ☐ | ☐ |
| Thoughts that you would be better off dead, or of hurting yourself in some way? | ☐ | ☐ | ☐ | ☐ |

Q46 In the **past month**, thinking about a stressful event related to the coronavirus outbreak, have you…(Please tick one box for each row)

| | Yes | No |
|---|---|---|
| had nightmares associated with your involvement with the event(s) or thought about your involvement with the event(s) when you did not want to? | ☐ | ☐ |
| tried hard not to think about your involvement with the event(s) or went out of your way to avoid situations that reminded you of your involvement with the event(s)? | ☐ | ☐ |
| been constantly on guard, watchful, or easily startled? | ☐ | ☐ |
| felt numb or detached from people, activities, or your surroundings? | ☐ | ☐ |
| felt guilty or unable to stop blaming yourself or others for your involvement with the coronavirus outbreak or any problems your involvement with the event(s) may have caused? | ☐ | ☐ |

Q47 Thinking over the **last two weeks**, how often have you been bothered by the following problems? (Please tick one box for each row)

| | Not at all | Several days | More than half the days | Nearly every day |
|---|---|---|---|---|
| Feeling nervous, anxious or on edge | ☐ | ☐ | ☐ | ☐ |
| Not being able to stop or control worrying | ☐ | ☐ | ☐ | ☐ |
| Worrying too much about different things | ☐ | ☐ | ☐ | ☐ |
| Trouble relaxing | ☐ | ☐ | ☐ | ☐ |
| Being so restless that it is hard to sit still | ☐ | ☐ | ☐ | ☐ |
| Becoming easily annoyed or irritable | ☐ | ☐ | ☐ | ☐ |
| Feeling afraid as if something awful might happen | ☐ | ☐ | ☐ | ☐ |

Q48 Over the last two weeks, please indicate how angry you have been feeling about being told to "shield"?

☐ Not at all
☐ Somewhat
☐ Moderately
☐ Very much

### About you

Q49 What is your age?

- ☐ 18 to 24
- ☐ 25 to 44
- ☐ 45 to 69
- ☐ 69+

Q50 Are you...

- ☐ Male
- ☐ Female
- ☐ Other
- ☐ Prefer not to say

Q51 To which of these groups do you consider you belong?

- ☐ White-British
- ☐ White-Irish
- ☐ White-other
- ☐ Chinese
- ☐ Asian
- ☐ Black or Black British
- ☐ African
- ☐ Mixed
- ☐ Other
- ☐ Would prefer not to say

Q52 Which of the following best describes your education level?

- ☐ Degree or above
- ☐ Below degree level
- ☐ Other
- ☐ None (no formal qualification)
- ☐ Would prefer not to say

Q53 Which of the following best describes your current employment status? (if you have more than one job, please select the option that best describes your **main** job, i.e. the job you do most often)

- ☐ Working full time (30 hours a week or more)
- ☐ Usually working full time (30 hours a week or more), but currently on leave or furloughed
- ☐ Working part-time (8-29 hours a week)
- ☐ Usually working part-time (8-29 hours a week), but currently on leave or furloughed
- ☐ Stay at home parent/homemaker/housewife or househusband
- ☐ Unemployed (registered or in process of registering)
- ☐ Unemployed (not registered but looking for work)
- ☐ Retired
- ☐ Student
- ☐ Don't know
- ☐ Would prefer not to say
- ☐ Other

If other, please provide details:

Q54 Please use this space to provide any additional comments about your experience of "shielding":

Thank you for your participation. The information you have shared will help us provide support for people who are "shielding".

If you have any queries, please

Please return your completed survey and consent form in the reply-paid envelope provided.
