## Supplement 2 for "Exploring the impact of shielding advice on the health and wellbeing of individuals identified as extremely vulnerable and advised to shield in Southwest England amid the COVID-19 pandemic: A mixed-methods evaluation"

**Self-isolation interview guide: Shielding group**
(Clinically extremely vulnerable people who have been advised to "shield")

**Introduction**
Thank you for agreeing to take part in this interview. Before we start, I'd just like to remind you that all information that you give will be confidential, and any published data from these interviews will be anonymous. I'd also like to remind you that I am recording this interview with a digital recorder – this recording will only be used to allow us to analyse the data collected and audio recordings will be used for research purposes only.

We are interviewing people who were advised to stay at home at all times and avoid all face to face contact with other people (known as "shielding"), because they are clinically extremely vulnerable to severe illness from coronavirus infection. We are trying to find out what it was like trying to do this, especially any difficulties you encountered, so we can provide advice and support for other people who have to "shield".

**Experiences of being advised to "shield"**
- **Can you start off by telling me about the first time you were contacted and advised that you needed to "shield"?**

*Prompts*
- *Were you already self-isolating?*
- *Were you aware you needed to "shield" before you were contacted?*

**Experiences of "shielding"**
- **How did you feel about being asked to do this?**

*Prompts*
- *How did you feel about being identified as someone who needed to "shield"*
- *Was there anything you were particularly concerned about?*

- **Did you make a decision about whether or not to do this?**
- *If not, why? [Then go to question about not following advice]*

**[For those who decided to shield]**
- **How did you go about trying to do this?**

*Prompts*
- *Can you tell me what steps you took at first?*
- *Has anything changed now that you have been doing it for a while?*

- **Can you tell me about your experiences of trying to follow the advice you were given?**

Prompts
- *Can you tell me what steps you took at first?*
- *Did anything change over time?*
- *Main problems with trying to self-isolate?*
- *Day to day living arrangements*
- *Access to basic supplies*
- *Access to basic care*
- *Family or other household members*
- *Work/study*
- *Finances*
- *Solutions to help with these problems?*

**[For those who decided NOT to shield]**

- **We realise that it may not be possible for everyone to follow all the advice – can you tell me about the times when it was not possible for you to follow advice that you have received?**

*Prompts*

- *Food supplies & medicine*
- *Work/livelihood*
- *Caring responsibilities*
- *Caring for animals*
- *Outdoor space*

- **Do you feel that advice to "shield" had an impact on your health and wellbeing in any way?**

*Prompts*

- *Were there any aspects that particularly affected you?*
- *Was there anything that affected your physical health?*
- *Was there anything that affected your mental health?*
- *Is there anything that might help make it easier for you?*

**Support**

- **What forms of support have you found you need during this time?**

*Prompts*

- *Practical support*
- *Social support*

- **What forms of support have you been able to get and who from?**

*Prompts*

- *Public Health England*
- *Other health services*
- *Government support service*
- *NHS volunteer responders*
- *Friends/family*
- *Neighbours*
- *Voluntary organisations*
- *Do you think there's any other support that might have been helpful?*

**Information and advice**

- **Did you get any information or advice from other places?**

*Prompts*

- *Internet*
- *Media*
- *Social media*
- *Friends/family*

- **What did you think of this information or advice?**

*Prompts*

- *Did any of it conflict with PHE guidance?*
- *What sources did you find most reliable?*

**Open comments**

- **Overall, what do you think of the advice you were provided?**
- **What would you want to tell someone who has just been asked or decided to "shield"?**
- **Do you think your experience has changed since the lockdown?**

- **How do you think your situation compares with people who are in lockdown?**
- **Is there anything else you would like us to know about your experience that we haven’t already covered?**

**Additional question – do not ask if long interview**

- **If a COVID vaccination was available for free on the NHS, how would you feel about it?**

- Would you have it?
- Why/why not?

Voucher
Debrief
Address or email
